# Prevalence of facemask use among general public when visiting wet market during Covid-19 pandemic: An observational study

**DOI:** 10.1101/2020.05.17.20105023

**Authors:** Gobi Hariyanayagam Gunasekaran, Sera Selvanthansundram Gunasekaran, Shargunan Selvanthan Gunasekaran, Nur Syafina Insyirah Binti Zaimi, Nor Amirah Binti Abdul Halim

**Affiliations:** Oncology Pharmacy, Hospital Seri Manjung, 32040 Seri Manjung, Perak, Malaysia; Medical Officer, Hospital Seri Manjung, 32040 Seri Manjung, Perak, Malaysia; Dental Officer, Manjung District Dental Clinic, 32000 Sitiawan, Perak, Malaysia

**Keywords:** Covid-19, personal protective equipment, facemask, odds ratio, Malaysia

## Abstract

**Background:** In late December, 2019, an outbreak of a novel coronavirus disease (COVID-19; previously known as 2019-nCoV) was epidemiologically linked to a seafood and wet animal wholesale market, in Wuhan, Hubei, China. This have instigated stigma among general population as wet market is viewed as high risk location for getting infected with coronavirus.

**Objective:** This study investigated the prevalence of facemask use among general population visiting wet market. This study also investigated the demographic factors contributing to unacceptable facemask practice.

**Setting:** This prospective observational study was done among visitor to a district wet market selling range of live or freshly slaughtered animals during COVID-19 pandemic outbreak.

**Methods:** Individuals entering through dedicated entry point were observed for the type, category and practice of wearing personal protective equipment. Inclusion criteria for this study were any individual’s entering the wet market. Subjects were categorized into two groups of acceptable and unacceptable facemask practice. The Pearson chi-square was used to test for differences in investigated variables in the univariate setting and Binary Logistic regression model was used in the multivariate setting.

**Main outcome measure:** Prevalence, acceptance practice and odds ratio of unacceptance of facemask use.

**Results:** Among 1697 individuals included in the final analysis, 1687 (99.7%) was observed wearing facemask with 1338 (78.8%) using medical-grade facemask. Among them, 1615 (95.7%) individuals’ facemask practice was acceptable while the reaming 72 (4.3%) individuals was observed with unacceptable facemask practice. Individuals using medical grade facemask and high-risk age group are 6.4 times (OR=6.40; 95% CI, 2.00-20.43; p=. 002) and 2.06 times practice (OR=2.06; 95% CI, 1.08-3.94; p=.028). More likely to practice unacceptable facemask use respectively.

**Conclusion:** High saturation of facemask among general population is an adequate indicator of public hygiene measures strategy which can help to mitigate the COVID-19 epidemic impact. Alarmingly, the unacceptable facemask practice among high-risk population raises the need for a targeted approach by healthcare authorities to ensure satisfactory facemask use.

## Introduction

In late December 2019, pneumonia cases of an unknown etiology was reported in Wuhan, China[1, 2]. The patients initially presented with acute systemic and respiratory disorders with clinical presentations resembling viral pneumonia [3–6] with 1.5-3.6% fatality rate [7, 8]. The pathogen of the outbreak was later identified as a novel beta-coronavirus, named COVID-19 (previously known as 2019-nCoV or SARS-CoV-2).

However, Covid-19 was not the first incidence of zoonotic pathogenic outbreak in wet markets as there have been several other coronaviruses outbreak within the past two decades. During 2002-2003, SARS (severe acute respiratory syndrome) outbreak originating from a wet market in Guangdong province of China was reported. Altogether, 8,422 cases of SARS with 916 deaths across 29 countries with a case fatality of 11%) [9]. In 2009 the influenza (H1N1) pandemic spread to 214 countries and caused an estimated 500,000 deaths with a case fatality rate of around 0.2%[10]. Similarly, coronavirus was responsible for the Middle Eastern Respiratory Syndrome (MERS), a severe respiratory disease outbreak in 2012 originating from Saudi Arabia. Likewise, There were 2,494 confirmed cases with 858 fatalities with case fatality rate of 34.4% [11]

The china health authorities were more prepared to handle outbreak this time around as they were able to identify and diagnose novel pathogens within 4 weeks from the first patient being identified compared to 4 months it took to alert the WHO during the 2003-2003 SARS outbreak [12]. The identification of a novel coronavirus outbreak was quickly followed by the closure of the Hunan market on January 1, 2020, after an epidemiologic alert by the local health authority due detection of 66% positive COVID-19 cases among the stall operators [3, 13] with the aim of preventing any further zoonotic transmission. This was in contrast during 2003-2003 SARS outbreak there was a delay in identifying the civet as a reservoir for the disease and civets continued to be sold on food markets. (SARS-CoV-2 shares 79% sequence identity with SARS-CoV) [14, 15]. Initial Public health strategy such as closing the of wet market have been frequently exercised to prevent further transmission of the disease [16, 17]. During the early stage of the outbreak; graphic pictures of civilian, authorities and health care personnel wearing extensive personal protective equipment (PPE) were widely covered by media highlighting the importance hygiene barriers in preventing infection[18]. Coupled with reports of the disease originating from wet market and COVID-19 transmission occurs through respiratory droplets from coughing and sneezing[19, 20], the use of personal protective equipment when visiting wet market is seen as vital step in reducing human-to-human transmission

However, in this unprecedented worldwide pandemic, the sociodemographics usage of facemask among general population is relatively unknown[21]. Investigating the prevalence of facemask use among general population when visiting wet market is a good indicator of social adaptability in response to local disease outbreak. The findings of this research could be used to improve strategic management for public health as well as managing Covid-19 pandemic in community setting.

### Aim of the study

This study aims to investigate the prevalence and types of respiratory protective device (facemask) usage among individuals visiting wet market during Covid-19 pandemic.This study also aims to evaluate the acceptance of the facemask practice worn by individuals.

### Ethics Approval

The ethical approval to conduct the study was obtained from the Medical Ethical Review Committee [KKM/NIHSEC/P20-1002(6)] Ministry of Health, Malaysia.

## Methods

### Study setting

This prospective observational study was conducted among individuals visiting a prominent wet market in Sitiawan, Perak, Malaysia in April 2020. During Covid-19 pandemic, the city council have closed the all peripheral entrance to the market and customers could only enter the market via the main entrance. Additionally, City council worker are station at the entrance to ensure social distancing and control the movement of individual entering and exiting the market through dedicated entry and exiting points. At the time of writing, the operating hour of the market was restricted from 6 am to 12 pm. The required data was recorded based on observation by data collectors who were stationed at strategic entry point.

### Inclusion and Exclusion

Inclusion criteria for this study were any individuals entering the wet market from selected entry points. Exclusion criteria for this study were Individual identified as stall operators, Health department worker, City council worker, individuals which are suspected of multiple entry and individuals who are exiting the treatment facility entrance.

### Data Variables

Individual data were collected by visually observing the type of facemask used and evaluating the garbing practice among visitors entering into the treatment facility. The following demographic data were collected: patient’s gender, age group and ethnicity while facemask data such as category and type of the product as well as garbing technique was recorded. Besides, the time of entrance to the facility was recorded. Gender was categorised as either male or female while individuals ethnicity was categorised into Malay or Non-Malay to reflect population distribution[22]. The Visitors age group was recorded as either as children, adult or elderly which was done based on subject’s facial and physical feature[23]. The age group was further categorised to low-risk age (children and adult) or high-risk age (elderly) group [24–26]. Facemask usage classifies as either “Yes” when any type of respiratory protective device is worn or as “No” when the product is absent. The category of facemask used was described according to their class; surgical facemask (2, 3 ply or any medical grade mask), respirators (all respirator Standard; FFP1 & P1, FFP2 & P2, N95, N99 & FFP3, P3, N100), cloth or paper mask. The facemask was further categorized as medical-use (Surgical facemask and respirator) or non-medical use (cloth and paper mask).The acceptance level of facemask practice was recorded as acceptable (correct wearable method) or unacceptable (incorrect method). The reason for unacceptable practice was further described as well.

### Statistical analysis

All demographic and categorical variables were presented as number (n) and percentage (%). Pearson’s chi-squared test was used to determine the statistically significant difference between the demographic characteristic between age group and the acceptance level of facemask practice. Simple logistic regression was used to screen the independent variable. Variables with p value <0.25 were included in the multivariate analysis. Binomial logistic regression test was applied to determine the contributing factor to unacceptable facemask garbing practice. Correlation matrix was checked for interaction between the variables. The Hosmer and Lemeshow test and Classification table was used to evaluate the model of good fit. The final model was presented with 95% confidence interval (CI) and its corresponding p-value. For all test Two-tailed p-value <0.05 was considered as statistically significant. All statistical analyses were performed using SPSS for Windows version 22.0 (SPSS Inc., Chicago, Illinois, USA).

## Result

The 1697 individuals included in the final analysis compromised of 60.3% (1024) male and 39.7% (673) female subjects with majority of 68.8% (1168) representation of Malay ethnic as. As shown in Table 1, we observed high saturation of face mask usage as 1687 (99.7%) of individuals had worn facemask. Among them 1338 (78.8%) individuals worn medical grade facemask with a majority of them was wearing surgical type facemask (76.5%).

**Table 1.** Demographic characteristic, facemask type and usage among general population during visit to wet market

| Description | Frequency (n=1697) | Percentage (%) |
| --- | --- | --- |
| **Gender** |  |  |
| Male | 1024 | 60.3 |
| Female | 673 | 39.7 |
| **Ethnic** |  |  |
| **Malay** |  |  |
| Malay | 1168 | 68.8 |
| **Non-Malay** |  |  |
| Chinese | 125 | 7.4 |
| Indian | 382 | 22.5 |
| Unidentifiable | 22 | 1.3 |
| **Age Group** |  |  |
| **Low-risk Age group** |  |  |
| Children | 5 | 0.3 |
| Adult | 1546 | 91.1 |
| **High-risk Age group** |  |  |
| Elderly | 146 | 8.6 |
| **Facemask Use** |  |  |
| Yes | 1687 | 99.7 |
| No | 10 | 0.6 |
| **Category of Facemask** |  |  |
| Did not wear face mask | 10 | 0.6 |
| **Medical Grade** |  |  |
| Surgical facemask | 1299 | 76.5 |
| Respirator | 39 | 2.3 |
| **Non-Medical Grade** |  |  |
| Cloth mask | 258 | 15.2 |
| Paper mask | 91 | 5.4 |

One of the main aim of the study was to investigate the prevalence of facemask practice among high-risk age group. A significant relation was found between age group and demographic variables of gender and use of facemask. As shown in Table 2, higher proportion of 117 male individuals (11.4%) were visiting wet market compared to 29 female individuals (4.3%) from high-risk age group, χ2(1) = 26.16, p <0.001. As for the use of facemask, equal proportion (50%) of individuals without facemask was reported between both age group. however, the proportion of facemask use within low-risk age group (99.7%) was higher compared to high-risk age group (96.6%), χ2(1) = 21.93 p <0.001. on the Contrary, no significance difference were observed among ethic and category of facemask between low-risk and high-risk age group.

**Table 2.** Demographic characteristic between low-risk and high-risk age group (n=1697)

|  |  |  |  |  |  |
| --- | --- | --- | --- | --- | --- |
| Description | Low-risk (n=1551) |  | High-risk (n=146) |  | p-value |
|  | Frequency | Percentage (%) | Frequency | Percentage (%) |  |
| **Gender** |  |  |  |  | <.001 |
| Male | 907 | 88.6 | 117 | 11.4 |  |
| Female | 644 | 95.7 | 29 | 4.3 |  |
| **Ethnic** |  |  |  |  | .780 |
| Malay | 1069 | 91.5 | 99 | 8.5 |  |
| Non-Malay | 482 | 91.1 | 47 | 8.9 |  |
| **Face Mask Use** |  |  |  |  | <.001 |
| Yes | 1546 | 91.6 | 141 | 8.4 |  |
| No | 5 | 50.0 | 5 | 50.0 |  |
| **Face mask category** |  |  |  |  | .082 |
| Medical Grade | 1218 | 91.0 | 120 | 9.0 |  |
| Non-Medical Grade | 328 | 94.0 | 21 | 6.0 |  |

The acceptance level were analysed between individuals who have worn facemask. As shown in Table 3, within 1687 individual who worn facemask, 1615 (95.7%) individuals’ facemask practice was acceptable while the reaming 72 (4.3%) subject was observed with unacceptable facemask practice. A significant relationship was found between facemask use and the variable of category of facemask and age group. Higher proportion of unacceptable facemask use was observed among 72 individuals (5.4%) using medical grade facemask compared to 3 individuals (0.9%) using non-medical grade face mask, χ2 (1) = 13.321, p<.001. As for age group, higher proportion of 12 individuals (8.5%) had unacceptable facemask use compared to 63 individuals (4.1%) from low-risk age group, χ2 (1) = 5.984, p=.029. Within 75 subjects with unacceptable facemask practice, 58 was wearing the wrong side out and the remaining 17 wore the mask loosely exposing either the nose, mouth or both while.

**Table 3.** Demographic characteristic between acceptance of facemask use (n=1687)

|  |  |  |  |  |  |
| --- | --- | --- | --- | --- | --- |
| Description | Acceptable (n=1615) |  | Unacceptable (n=72) |  | p-value |
|  | Frequency | Percentage (%) | Frequency | Percentage (%) |  |
| **Gender** |  |  |  |  | .070 |
| Male | 963 | 94.8 | 53 | 5.2 |  |
| Female | 649 | 96.7 | 22 | 3.3 |  |
| **Ethnic** |  |  |  |  | .308 |
| Malay | 1105 | 95.2 | 56 | 4.8 |  |
| Non-Malay | 507 | 96.2 | 19 | 3.6 |  |
| **Category of face mask** |  |  |  |  | <.001 |
| Medical Grade | 1266 | 94.6 | 72 | 5.4 |  |
| Non-Medical Grade | 346 | 99.1 | 3 | 0.9 |  |
| **Age group** |  |  |  |  | .029 |
| Low-Risk | 1483 | 95.9 | 63 | 4.1 |  |
| High-Risk | 129 | 91.5 | 12 | 8.5 |  |

Simple logistic regression was used to screen demographic variable which contributed to unacceptable facemask practice. To reflect the risk of morbidity from Covid-19 infection, demographic profiles with low mortality rate such as female, non-Malay, low-risk age group, non-medical grade facemask was chooses as the reference. Non-medical facemask was chosen as the reference due to the lower proportion (0.9%) of unacceptable practice the result are presented on Table 4.

**Table 4.**
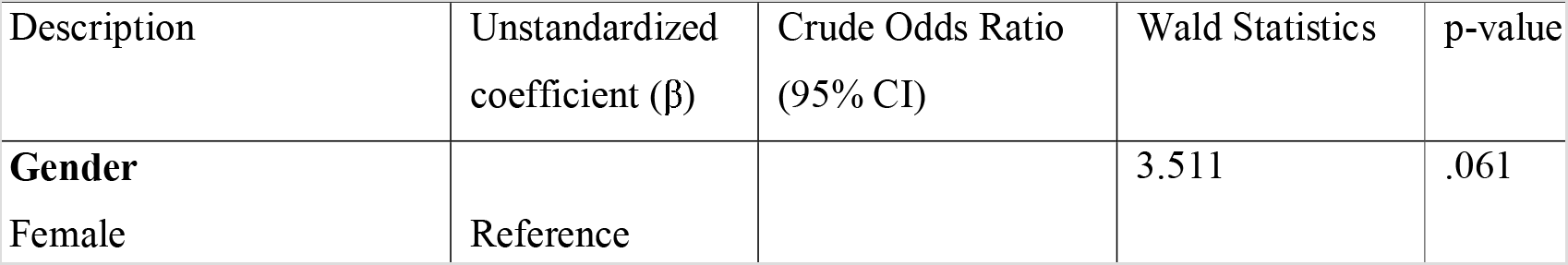
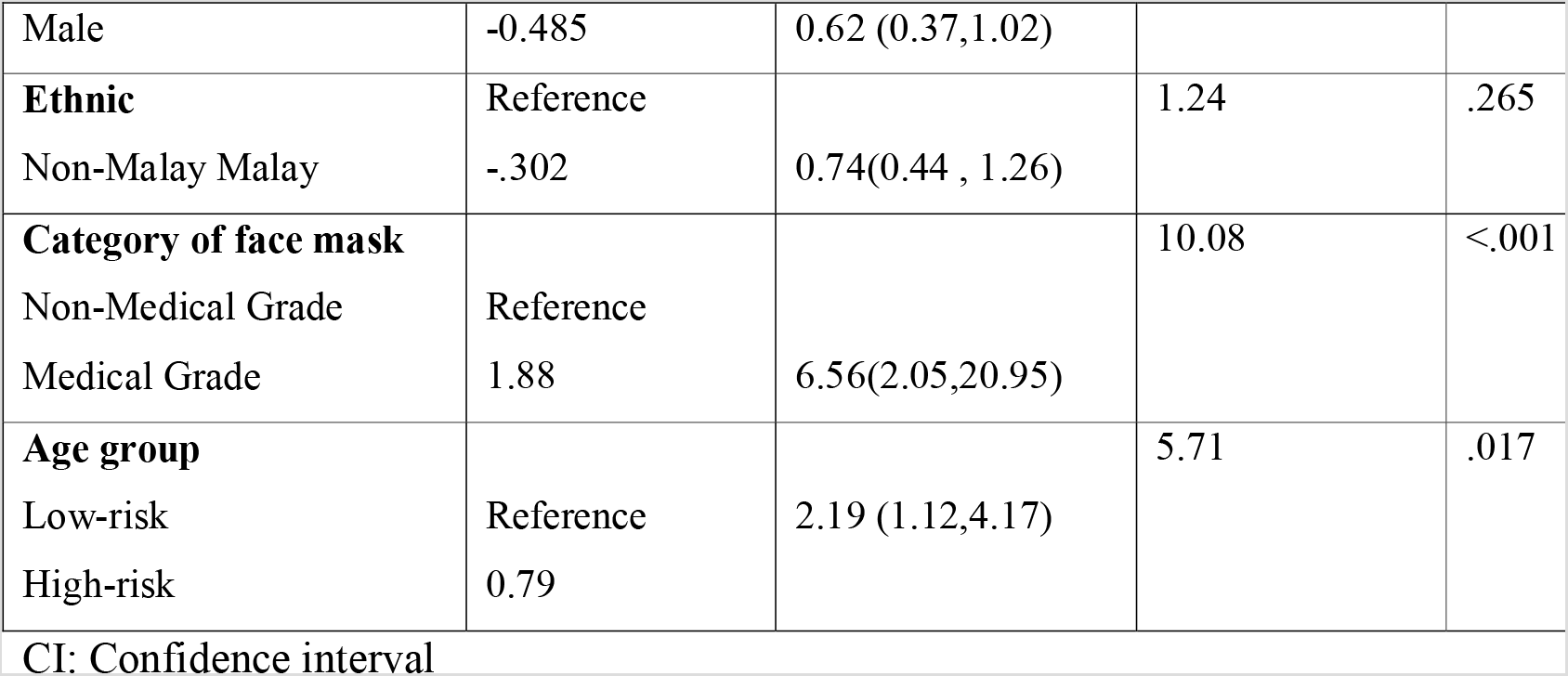
Associated factor of unacceptable facemask practice by simple logistic regression (n=1687)

Individuals wearing medical grade facemask are 6.56 times more likely to practice unacceptable facemask practice (Odds Ratio; OR=6.56; 95% CI, 2.05-20.95; p<.001). Subsequently, high-risk age group individual are 2.19 times more likely to practice unacceptable facemask practice (OR=2.19; 95% CI, 1.12-4.17; p=0.17).

As shown in Table 5, the final binary logistic regression model for unacceptable facemask garbing practice was adjusted for gender, ethnic and age group.

**Table 5.**
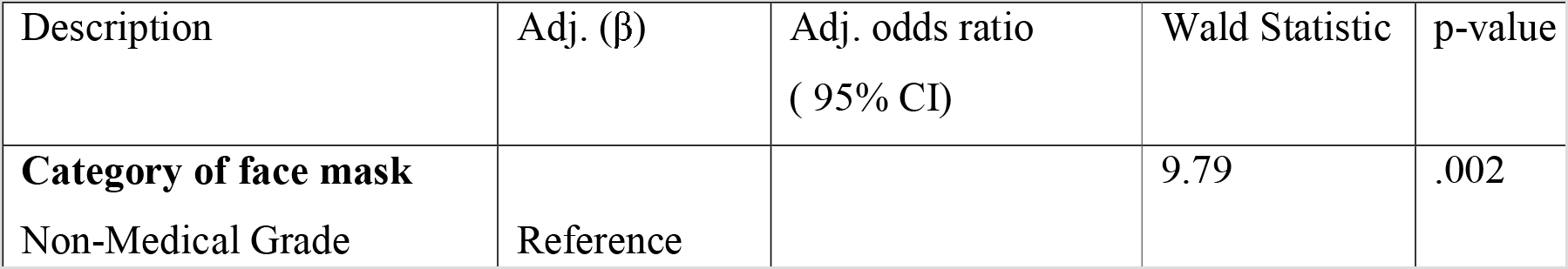
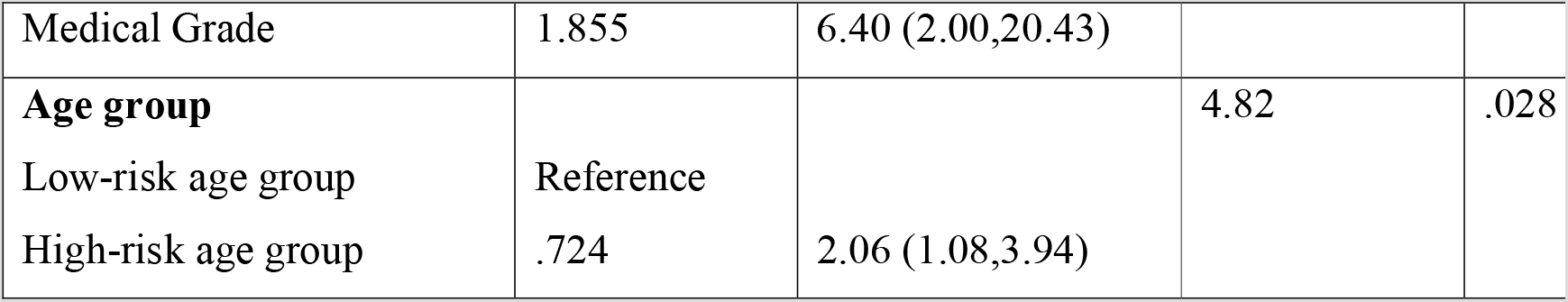
The adjusted factor of unacceptable facemask practice by Multiple logistic regression

Adj.: Adjusted Regression coefficient, Adj. odds ratio: Adjusted odds ratio, CI: Confidence interval. The final model was obtained using backward linear regression model. Correlation matrix was checked for interaction between the variables was small. The 2-way interaction between the categorical was not significant. The Hosmer and Lemeshow test significant was ϰ^2^ (5) = 2.488, p =.778 while model summary -2 Log likelihood= 588.069 and Classification table (overall classification percentage 95.6%) indicating the model fit.

Similarly, adjusted for other variables, individuals using medical grade facemask had 6.4 times more likely to practice unacceptable facemask garbing (OR=6.40; 95% CI, 2.00-20.43; p=.002). Similarly, high-risk age group individuals are 2.06 times more likely to practice unacceptable facemask practice (OR=2.06; 95% CI, 1.08-3.94; p=.028).

## Discussion

COVID-19 pandemic is the latest among string of zoonotic origin outbreak in the modern time.To date, most efforts have focused on clinical management, defining the spectrum of disease and tracking morbidity and mortality of SARS-CoV-2 infection [27, 28]. As no effective treatment is available, health care authorities have relied on public health management to mitigate local human-to-human transmission. Initial recommendation of worldwide health authorise have generally been along the line of practicing social distancing and using personal protective equipment’s [29, 30].

As such, the high prevalence of facemask use among visitors to the wet market (99.7%) observed in our study is similar to another local study among visitors to hospital (96.9%) [31], indicating high social adaptability general population in response to local disease outbreak. the larger proportion of facemask use among visitors to wet market could be contributed by widespread public stigma where wet markets are viewed as a high risk location for potential infection[32, 33].

The high saturation of facemask usage should be welcomed as the rationale behind wearing facemasks has been widely discussed in relation to preventing human-to-human transmission[34, 35].Although the consensus of asymptomatic individuals transmitting the virus before symptoms develop seems to be inconclusive, a risk on transmission cannot be fully excluded [7, 36, 37]. While there was consistency in the recommendation that symptomatic individuals and health care workers should use facemasks, discrepancies in recommendations on the facemask use by general population use varied greatly between countries. Feng S et al and team compiled contradictory view by difference agencies; generally the western countries such as the United States, United Kingdom and Germany have stand use of facemask by healthy general population while Asian countries such as China, South Korea and Japan have adopted a risk-based approach by distributing facemask to the general public[38]. The different approach in facemask by Asian countries are the response adaptive and preventive behaviour due to previous exposure of regional SARS epidemic[39]. During the SARS epidemic, preventive measures such as face mask use were commonly practiced[40] and have been norm of the public even after the epidemic subsided [41].

It’s evident that the approach to facemask usage among general population depends on the recommendation of local health authorities as well as the availability of the commercial product in the market. In Malaysia, there was reports of facemask shortage during the initial outbreak[42]; however, the widespread use and availability of the facemask observed in this study could be due to importation on 10 million facemask from china during the acute shortage phase[43], increase in manufacturing and establishment of new manufacturing facility to increase in production capacity of local manufacturer [44, 45] and handling out 24.6 million facemask to Malaysian household[46].Despite the high prevalence of facemask usage among our population, we found that high-risk age group have unacceptable facemask practice. The unacceptable facemask practice among high-risk age group raises concern as subjects ≥ 60 years old are 18 times (OR: 18.82, 95% CI 7.20-41.55) more likely to die from COVID-19 [47] and multiple other studies have established mortality risk for this group [8, 48, 49].

Although the use of facemask should be encouraged, the use of medical grade facemask by general population is questionable. Acute shortage of facemask have been reported when local epidemics begin [50–52]. When China announced Hubei provenance lockdown, the daily demand of facemask estimated to be >50 million pieces daily whereas the daily production have dropped from 20 million to 15 million[53] promoting China to temporarily suspend the export of facemask. These have resulted in worldwide shortage of medical facemasks [50–52, 54]. This sudden uptake in the use of medical-grade facemasks by the general public worsens the global shortage of facemasks, further depletes facemask availability to both health-care workers and susceptible individuals[55, 56]

We observed high percentage of medical grade facemask usage in both high-risk age group and low-risk age group. However, the high proportion among low-risk age group raises the question on the necessity of medical grade facemask use in community setting. Besides that, we also observed that individuals using medical grade facemask had 6.4 times more likely to practice unacceptable facemask use. As evidence suggests COVID-19 could be transmitted through droplet [57, 58], improper facemasks may be ineffective for prevention as they generally do not form a tight seal against the face skin and hence are not likely to be effective [59]. Additionally, improver use of facemask raises the concerns such practice could provide a false sense of security which in turn could neglect other means of risk reduction such as social distancing and hand washing[60]. Non-compliance to facemask practice such as loosely fitted facemask, exposing mouth and nose as observed by us have been reported as main concern in previous by other researches as well [47, 53, 61]; however the compliance could be improved through targeted public health education[62, 63].

Evidence that facemasks can protect against infections in general population widely debated [62–64]. In 3 randomised clinical trial, wearing a facemask have reduced odds of developing respiratory symptoms, 0.6% (OR 0.94, 95% CI 0.75 to 1.19, I2 29%, low certainty evidence)[65]. two community based retrospective case-control studies in Hong Kong and China during previous 2003 SARS-CoV-1 outbreak reported that use of medical grade facemasks (surgical masks in both studies) was associated with at least 60% lower odds of contracting SARS [66, 67]. The contradicting view of facemask usage are due to the lack of conclusive Research finding which need to be done during a pandemic when facemask compliance is high enough for its effectiveness to be assured. However, a mathematical simulation model by Eikenberry, Steffen E., et al. suggest use of facemasks by the general public could potentially restrain community transmission and reduce mortality rate due to Covid-19 pandemic by 24–65% [68]. On the hind sight, the effectiveness of facemask usage in preventing human-to-human transmission could be evaluated by longitudinal study during this pandemic session.

While waiting for effective antiviral treatment against Covid-19, increasing evidence[69] and analysis[64, 70] supports the use of facemask as low-cost addition with social distancing and hand hygiene during the COVID-19 pandemic [63, 71]. Although the high saturation of face mask usage is welcomed, the mental wellbeing contributing to such high saturation of usage should not be neglected [72]. Lin et al. correlated all-time high search for “face mask” in Google could be a sign of anxiety appearing in the society[73]. Szczesniak et al. and teams findings imply that in addition of protection against the COVID-19, the use of facemask also increase the level of perceived self-protection and of social solidarity which, thereby improve mental health wellbeing[74].

The finding of our study are limited by the sociogeographic of study location. The availability and price of facemask on market could have greatly influenced subject’s preference. In addition, our population consist of individuals visiting wet market which is generally considered as high risk area for human-to-human transmission and hence visitors could have taken extra precaution which otherwise will not be practices could have skewed our observation. Besides that, we only observed gross facemask usage and were not able to assess neither the quality nor the fitting adequacy of the facemask used.

## Conclusion

In conclusion, Covid-19 infection zoonotic disease transmitted via droplets with no effective treatment is available at the time of writing. Hence, Public health management is paramount to mitigate local human-to-human transmission to reduce the stress on health care system. In spite of contradicting opinion on the potential value of facemasks for general population use; the widespread availability and lack of obvious harm in using facemask together with other environmental hygiene measures is a vital epidemiology strategy which may help to alleviate the COVID-19 epidemic impact.

## Data Availability

The datasets generated during and/or analysed during the current study are available from the corresponding author on reasonable request

## Acknowledgement

Our study group would like to thank Director General, Ministry of Health Malaysia for approval to publish this research.

